# Applying Fast Healthcare Interoperability Resources (FHIR) for Pathogen Genomics at the Point of Care

**DOI:** 10.1101/2024.03.10.24303884

**Authors:** Soyean Kim, Gordon Ritchie, Mahdi Mobini, Aishwarya Sridhar, Joseph Amlung, Andrew S. Kanter, Bryn Rhodes, Robert H. Dolin, Bret S. E. Heale, William W. L. Hsiao

## Abstract

Modern-day microbial diagnostics and genomics have the potential to revolutionize individual and population-level infectious disease prevention, patient care, and treatment. To realize the potential, we need new approaches to standardizing testing and genomic data so that complex data and knowledge can be integrated at the point of care reliably and without ambiguity. We provide a series of approaches to pathogen genomic information standardization and guides to improve data interoperability which is key to harnessing the power of modern testing and genomics data.

We develop generalizable knowledge and guidance to integrate the systems of terminology management, data modeling and inference that can provide flexibility for collaborative development across multiple domains (i.e. public health, clinical, academic research and open source communities) in order to significantly speed up the applications of pathogen genomic knowledge.

We observed numerous advantages to using healthcare-specific standards such as Fast Healthcare Interoperability Resources (FHIR) and Clinical Quality Language (CQL). Advantages include convenient information models, mechanisms for verification, and the availability of tools, documentation and expertise to provide assistance during development. We also found the critical role of community-driven domain-specific ontologies which provide a source of terminologies thereby addressing content coverage gaps in the common clinical terminologies.

**Strengths and limitations of this study:** To our knowledge, this is the first work of its kind to provide structured guidance on pathogen genomic data interoperability using HL7 FHIR resources for a clinical scenario involving whole genome sequencing. We believe this provides a clear path for broader stakeholders including implementors and knowledge curators on how to collaborate and facilitate automation in support of speedy exchange of complex knowledge for genomic epidemiology.

We believe the tools and documentation provided can be a resource for clinical informatics, researchers, and public health organizations who want to collaborate, grow and exchange pathogen genomic knowledge for critical public health applications.

We acknowledge the limitations of this work.

First, the tools developed here are limited in scope and not yet validated among the broader FHIR community. Therefore the ability to generalize for a broad set of pathogens is limited. Standardization of external ontology will require approval from the HL7 terminology authority. This approval process will require the demonstration of quality processes and measures and licensing and legal processes as well as community buy-ins.

The information model here is developed based on scenario modelling. Additional validation using real clinical scenarios and patient data will be required for future developments. As the whole genome sequencing process is only beginning to emerge in clinical practices, more patient-derived whole genome sequence result data from multiple facilities will be needed to create generalized clinically valid pathogen genomic tools.

The privacy issues surrounding the utilization of social determinants of health data (SDOH), while taking into account the relational and structural aspects of infectious disease outbreaks that impact vulnerable communities, will further require careful consideration prior to standardizing the discovery and access of SDOH data.

**Preamble:** Modern-day medical diagnostics using microbial genomics have the potential to revolutionize individual and population-level disease prevention, patient care, and treatment. Clinical laboratories are increasingly pursuing pathogen genomics for infectious disease diagnosis and characterizing whole genome sequences of cultured isolates to help with infection prevention and control practices (IPAC) regarding outbreaks and surveillance

However, to achieve that goal, we need to consider the speed, complexity, and ability to integrate the point-of-care data with genomic data. We provide a series of approaches to pathogen genomic information standardization and guides to improve data interoperability, which is key to harnessing the power of modern testing and genomics data.

## Introduction

Clinical laboratories are increasingly pursuing pathogen genomics for infectious disease diagnosis and characterizing whole genome sequences of cultured isolates to help with infection prevention and control practices (IPAC) regarding outbreaks and surveillance (Cameron et al. 2020). For example, the presence of plasmid-mediated antimicrobial resistance (AMR) genes is an important concern because plasmid-mediated AMR genes can easily spread to other bacterial pathogens (Das 2023). Global crisis involving AMR rates outpacing the development of new antibiotics suggests this trend could result in common infections becoming difficult to treat or even untreatable, leading to longer treatment periods, longer hospital stays, higher medical costs, severe illness and death (Singh et al. 2022).

While implementing a generalizable, secure, and efficient computing infrastructure (van Heusden et al. 2022) and associated governance framework for real-time pathogen data exchange (Gardy, Loman, and Rambaut 2015) that would enable scientific research and data analysis remains a challenge, HL7 Fast Healthcare Interoperability Resources (FHIR) enables new possibilities to undertake speedy implementation of standard health care tools for rapid data sharing and semantic interoperability without ambiguity.

As the use of pathogen genomic analysis has become the standard for variant detection for SARS-CoV-2, more investigations are done, and new knowledge is created. As a result, we have new ontologies being developed and, therefore greater needs for terminology management. The growing needs for data management necessitate the ability to draw from clinical terminologies, such as SNOMED, as well as community-based open-source ontologies (e.g. Open Biological and Biomedical Ontologies (OBO) Foundry) that may overlap in domain knowledge. For example, in FHIR, each component entry needs a code. A set of concepts is assigned with specific codes to guide implementers in the correct use and understanding of the codes. A set of common codes organized as terminology standards is used. LOINC (Drenkhahn and Ingenerf 2020) and SNOMED (Lee et al. 2014) are some examples of common medical terminologies well adopted globally to describe most clinical operations. Yet, these code systems cannot curate every piece of knowledge in the growing field of genomics (Khorrami, Ahmadi, and Sheikhtaheri 2018). As new knowledge is created at an unprecedented pace, the curation and processing of such knowledge can no longer be contained within a single organization. Open-source ontology communities have a role in providing curated knowledge and available nomenclatures to supplement this knowledge curation gap. A community such as OBO Foundry supports ontology development and dissemination following FAIR Principles <u>(Wilkinson et al. 2016)</u>. FAIR data are data which meet principles of findability, accessibility, interoperability, and reusability (FAIR). By combining community-driven innovation with standardized ontology development, which includes a controlled vocabulary (comprising concepts, relations, instances, and axioms), we are poised to realize the benefits of this intricate knowledge more rapidly.

Consider a scenario where there will soon be standardized and routine pathogen genomic investigations within the hospitals. This would mean real-time clinical decision support (CDS) where bioinformatics computation can draw from massive genomic information as well as both broader curated knowledge database systems <u>(Cameron et al. 2020)</u>. Logic (for electronic clinical quality measurement and clinical decision support) and FHIR operations are two relevant areas that provide foundations for clinical decision support based on genomic data. Current efforts on EHR-genomics integration (Alterovitz et al. 2020) include a standardized suite of genomics operations via Application Programming Interface (APIs) for handling individual’s generic data to obviate the need for developers to simplify the required application development effort to bring contextually relevant genomic findings and recommendations to clinicians at the point of care (Dolin et al. 2023).

The purpose of this paper is to propose and evaluate a proof of concept FHIR-based system designed for pathogen genomic data involving a local bacterial outbreak scenario. We develop generalizable informatics tools (FHIR artifacts) and guidance to integrate the systems of terminology management, data modelling, and inference that can provide flexibility for collaborative development across multiple domains (i.e. public health, clinical, academic research, and open-source communities) while allowing for core FHIR capability for exchange to significantly speed up the applications of complex genomic knowledge.

## Methods

We investigated relevant terminology resources, FHIR resources and FHIR data models and model inference as the key components for pathogen genomic data interoperability to provide guidance for well-integrated systems and genomic integrated health care. All terminology resources were generated as CodeSystem and ValueSet to represent data elements including data type, cardinality and descriptions where the basic structures contain narratives as the HTML representation of the CodeSystem and the ValueSet. This was possible with the use of Open Concept Lab (OCL) terminology service which supports HL7 FHIR v4.0.1 based on the Mobile Sharing Value Sets and Concept Maps (mSVCM) IHE Profile. ClinFHIR<u>(clinFHIR Launcher)</u> was used to generate FHIR resources based on the shigellosis clinical scenario to represent the data model, which is to bind with the aforementioned terminology resources. In evaluating the computable clinical knowledge, we used Clinical Quality Language (CQL), a high-level, domain-specific language focused on clinical quality and FHIR operations as they provide foundations for clinical decision support by building computable measures using FHIR resources. Test data was generated using a DiagonosticReport resource, which contains the code to recommend that the AMR gene found is plasmid-mediated (using the experimental CodeSystem and ValueSet published at OCL). Given the nature of the local outbreak, plasmid transmission was assumed. In building CQL based artifacts and consuming FHIR resources, a CQL library was created to support the decision support implementation. This experiment was tested by running the CQL library and was carried out using VSCode local runtime. A FHIR Implementation Guide (IG) was created as a result to allow users to leverage the FHIR publishing toolchain, distribution and documentation of the CQL artifacts.

## Results

### The terminology management component of pathogen genomic data systems

In FHIR, coded elements are standardized, with the option to bind to specific terminology to represent more granular data models. The terminology artifacts used are referred to as CodeSystem and ValueSet. For example, Table 1 lists the elements used in the Figure 5. Consider the “category” element in Table 1, which is used to define a code that classifies the diagnostic service that created the report. Cardinality (a minimum and maximum number of required appearances) is shown as “Mult.”.

**Table 1:**
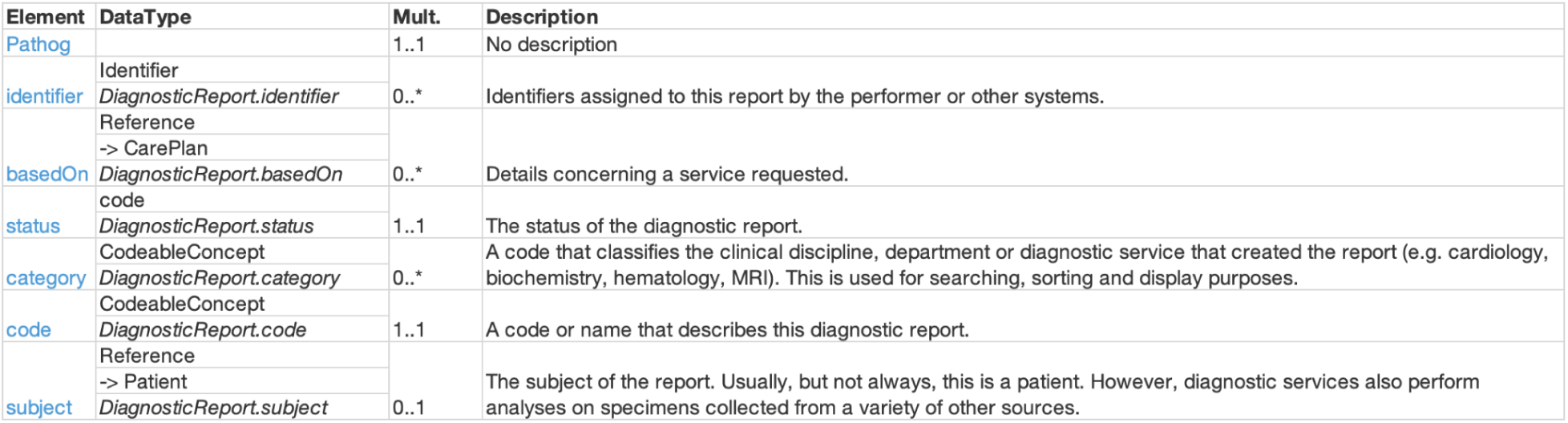
Terminology data elements and cardinality.

Electronic Health Record (EHR) systems use many codes from standardized sets of codes. From simple concepts such as patient’s sex to more complex ones (i.e. diagnoses, allergies and adverse reactions), FHIR cannot define every required code and related concepts for every clinical encounter across the health systems around the world. Instead, FHIR provides a mechanism to manage the codes and related concepts.

As FHIR’s CodeSystems and ValueSets are two key components of its terminology management systems, we closely examine the concepts behind CodeSystem and ValueSet and the FHIR resources that embody them for pathogen genomic use cases.

Figure 1 shows the basic structure of the resource. The top section like all FHIR resources contains a narrative as the HTML representation of the CodeSystem. The middle section is all the metadata about the CodeSystem, things like the version, a name and description for the CodeSystem, its status, whether it is experimental or not (experimental =”true”), who is the publisher (“cidgoh”) and whether the codes are case sensitive. The last section is the actual codes each in their own <concept> element showing the device type = “Oxford Nanopore GridION” in the experimental version of the GenEpiO CodeSystem.

**Figure 1:**
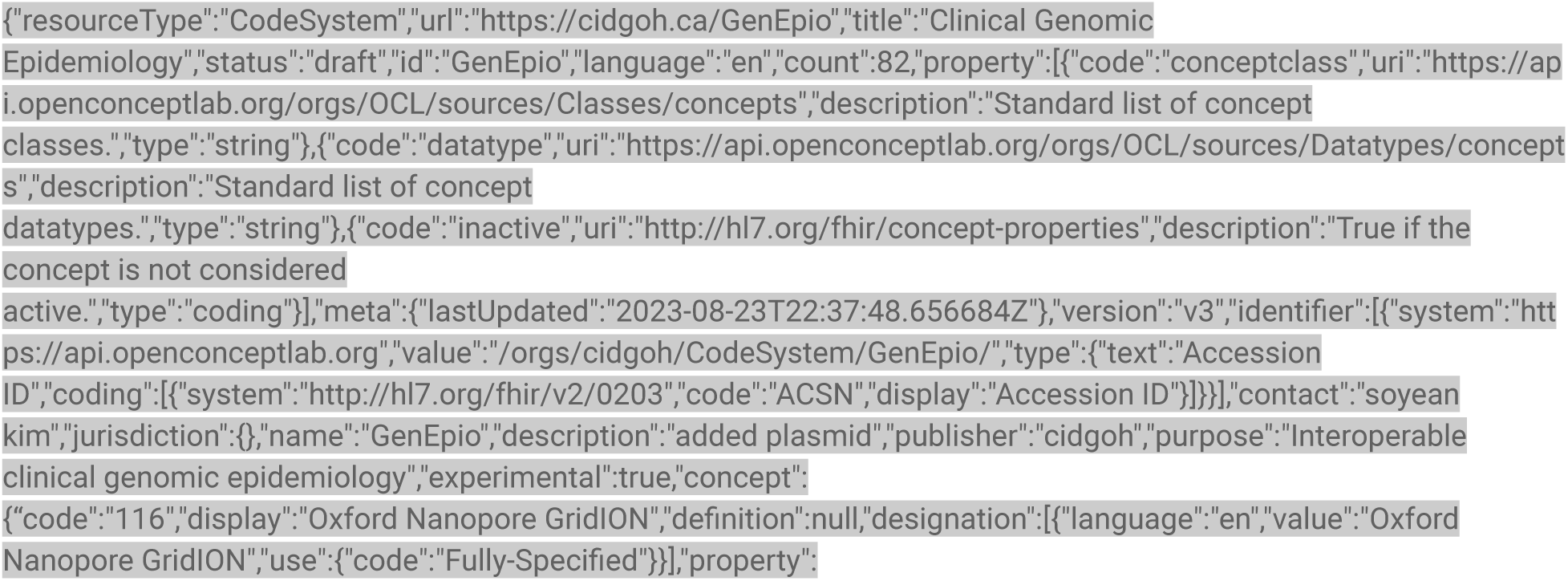
Experimental CodeSystem resource in Open Concept Lab (OCL)

ValueSet FHIR resourceValueSet specifies a set of codes drawn from one or more code systems, intended for use in a particular context.

Figure 2 showing ValueSet resource (ValueSet “ShigellaWGS”) selects codes from the CodeSystem example we looked at (the experimental version of the GenEpioO CodeSystem as hypothetical examples). Please notice how the CodeSystem is referenced in the ValueSet using the “compose include system” (line 6 in the JSON output) which points to the ‘url’ property /orgs/cidgoh/ValueSet/GenEpio/. This reference is a property of the CodeSystem resource. This ‘url’ is intended to be globally unique, so the same CodeSystem can be in any number of servers and can always be found by this ‘url’ property.

**Figure 2:**
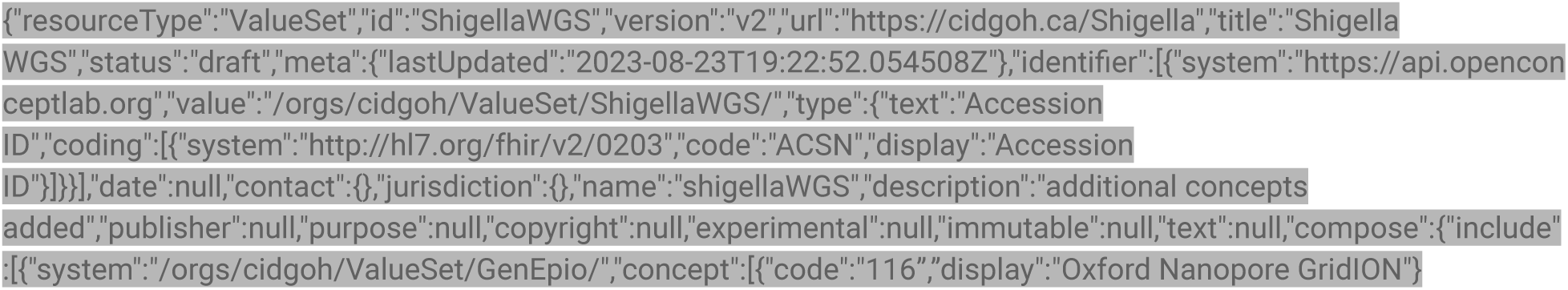
Experimental ValueSet resource

The ValueSet is free to pick and choose whichever codes it requires from the CodeSystem. In addition, it can select more codes from another CodeSystem if required.

### Terminology binding for pathogen genomic information model

Terminology binding provides a linkage between the information model and terminology and is an important part of supporting data collection, query, and semantic interoperability (Aso et al. 2013). Terminology server provides a wide range of terminology management services, which may include code system lookup ($lookup) and validation of codes ($validate-code). Leading terminology servers such as OntoServer (Metke-Jimenez et al. 2018) and OCL (“https://openconceptlab.org/) provide FHIR native terminology services supporting a broad range of SNOMED CT and LOINC features. We used OCL (https://sso.openconceptlab.org) to provide real-time electronic access to terminological resources to help users collaboratively manage, publish, and use terminology. One can easily create a repository of mixed ontology content using OCL. This provides opportunities for broader ontology communities (both open source and proprietary) to interoperate using FHIR Terminology Service capabilities (https://openconceptlab.org/terminology-service/). The experimental version (“GenEpiO”) is published at: https://app.openconceptlab.org/#/orgs/cidgoh/sources/

### Scenario modelling involving a clinical scenario for an individual patient

This clinical scenario is constructed to mimic the *Shigella* outbreak in Vancouver, BC, Canada based on contributions from the participating group and on-site meetings. The group involving clinical microbiologists, clinicians, and research teams met on multiple occasions between September 2022 and December 2023 and went through a series of scenario modelling exercises and discussed clinical scenarios involving whole genome sequencing (WGS). No personal identifiable information was used for this process. The discussion included the review of a sample report and applications of expanded pathogen genomics knowledge from WGS in daily clinical practice and how to best overcome the current limitations of clinical data sharing and use/integration of other apps and services. The group also discussed which FHIR tools can be best suited to the clinical scenario generated.

Consider the following scenario involving a patient who presented symptoms of diarrhea. The attending clinician is aware of the local *Shigella* outbreak and suspects shigellosis. The clinician orders a lab test that identifies *Shigella* bacteria in the stool of the patient based on a culture that isolates the bacteria. The clinician consults with the clinical microbiologist. The clinical microbiologist orders genotype testing using whole genome sequencing analysis to identify AMR genes and plasmids. Both tests confirm shigellosis. The WGS analysis found several AMR genes and detected plasmids. The AMR genes detected show chromosome intrinsic resistant markers. *Shigella* bacteria can spread in environments where there is crowding or access to clean water and toilets is limited. An increase in antimicrobial-resistant *Shigella* infections among people experiencing homelessness has been observed. Genotypes including the absence of tet (A) along with other characteristics <u>(Stefanovic et al. 2023)</u> were investigated for potential association with the homeless population.

A sample report was reviewed, which contains the analysis from the BugSeq pipeline (Fan, Huang, and Chorlton 2021). A screenshot of the sample report is shown in Figure 3 and Figure 4 for *Shigella sonnei*. The General Statistics section describes quality metrics for sequence coverage and assembly such as N50 and Assembly Length. The Plasmid Detection section in Figure 3 describes the identified plasmid (ClusterID), host range, and backbone information such as replicon and relaxease types part of plasmid reconstruction. The Genotypic Markers section in Figure 3 contains the names of antimicrobials, class of antibiotics and associated confidence in predicted antimicrobial resistance.

**Figure 3:**
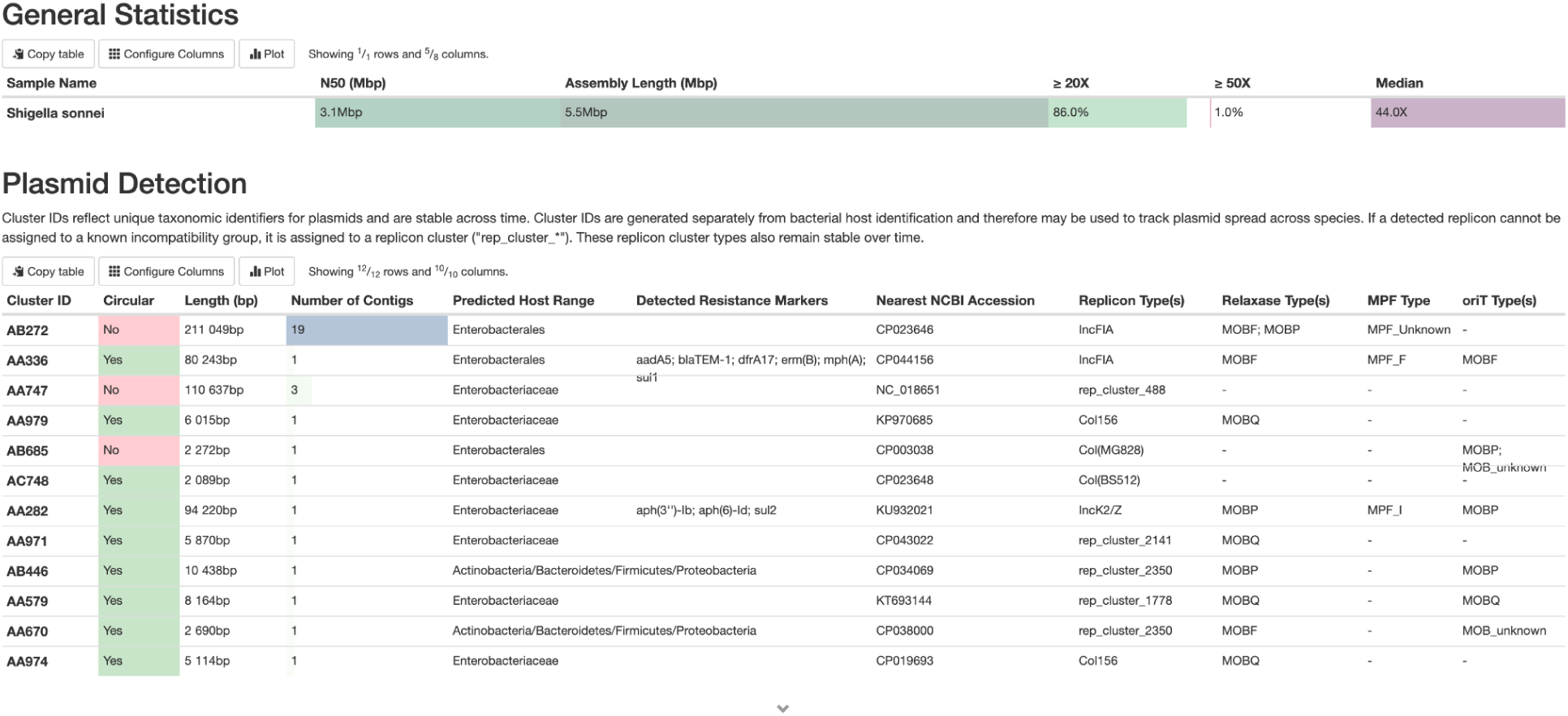
Sample output containing plasmid detection using BugSeq (bugseq.com V5.0 (Vancouver, Canada)

**Figure 4:**
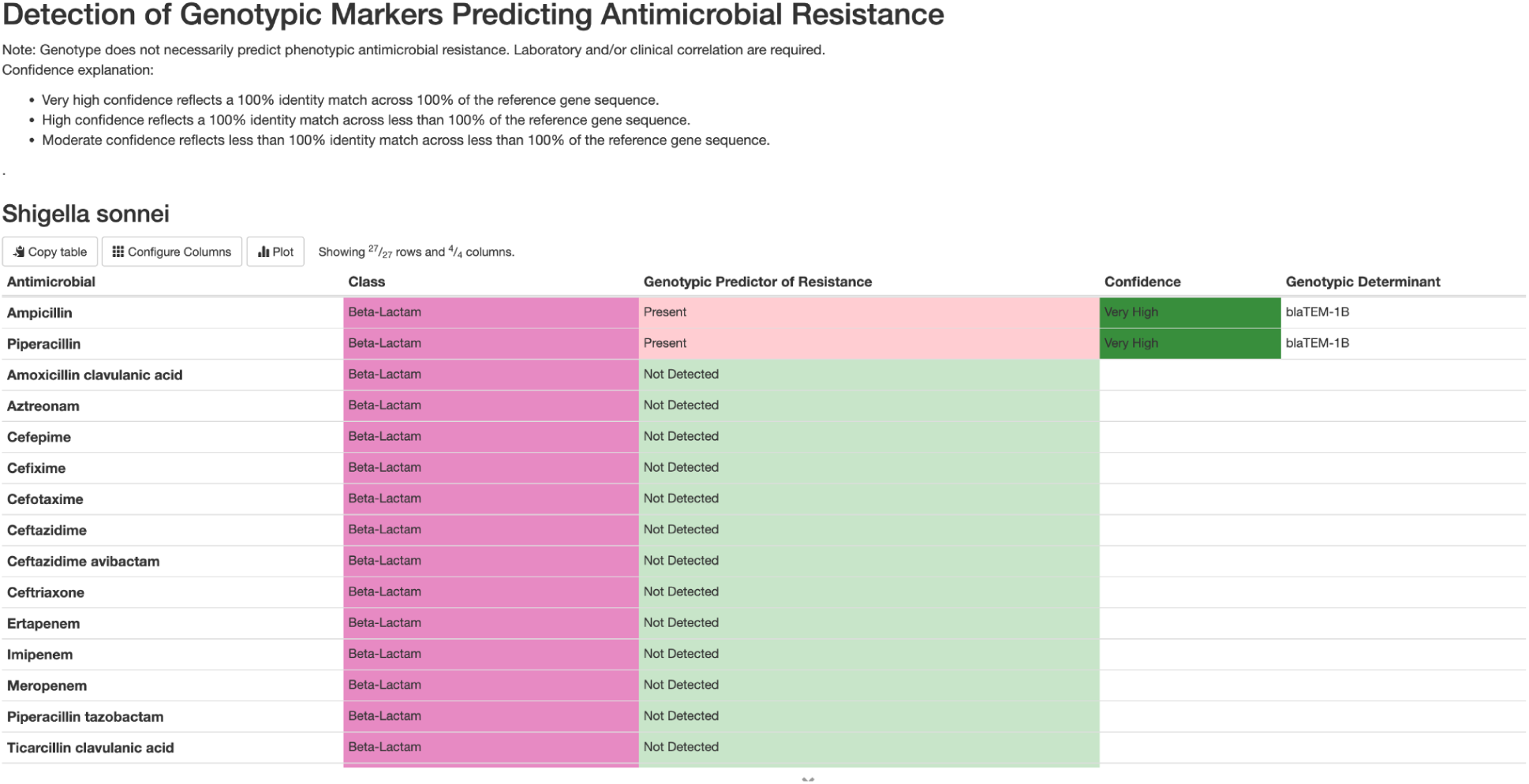
Sample output containing genotypic markers using BugSeq (bugseq.com V5.0 (Vancouver, Canada)

**Figure 5:**
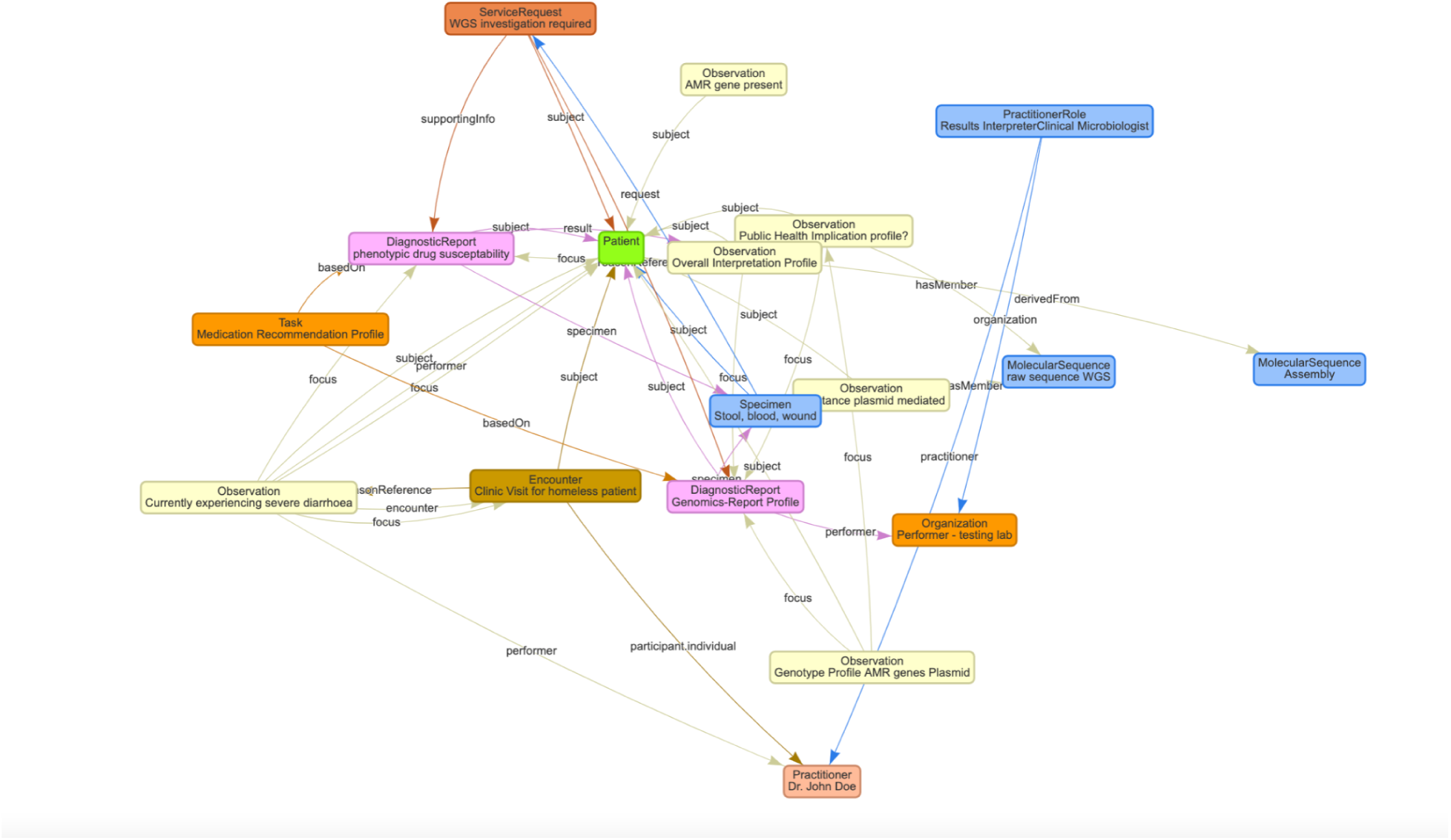
pathogen genomic data model

The Diagnostic Report resource for FHIR containing the genotypic information follows the simulated information model (see the data model component) that includes: 1) identifier, 2) details for the service requests for AMR gene detection and plasmid detection, 3) the status of the report, 4) service category to define the purpose, 5) code for the test to differentiate whole genome sequencing test for the organism from standard phenotypic lab tests, 6) the subject of the report as patient and organism, 7) describe the encounter when the test is ordered, 8) any clinically relevant date and time, 9) the practitioner or organization that is responsible for the report’s conclusions and interpretations including clinical microbiologist and/or bioinformaticians to assist interpretation of the whole genome sequence analysis findings, 10) describe the specimen this test is based on such as stool, blood and wound, 11) observation containing *Shigella* strains, detection of plasmid and plasmid information, 12) reference to imaging if any, and 13) clinical conclusions. The information model needs to bind with ValueSet from terminology servers. The results from the Diagnostic Report should reference an Observation resource. For more details of the Observation resource, please see the whole genome sequence analysis observation in the “Data model component”.

### The Data model component

By way of illustration, please see Figure 5 using FHIR resources designed based on the shigellosis clinical scenario described above. The following data model is published in ClinFHIR (https://clinfhir.com/).

The experimental CodeSystem described in Table 2 and Figure 6 is based on the aforementioned simulated clinical scenario involving *Shigella* whole genome sequencing analysis. Search and validation against the LOINC codes are performed using OCL.

**Figure 6:**
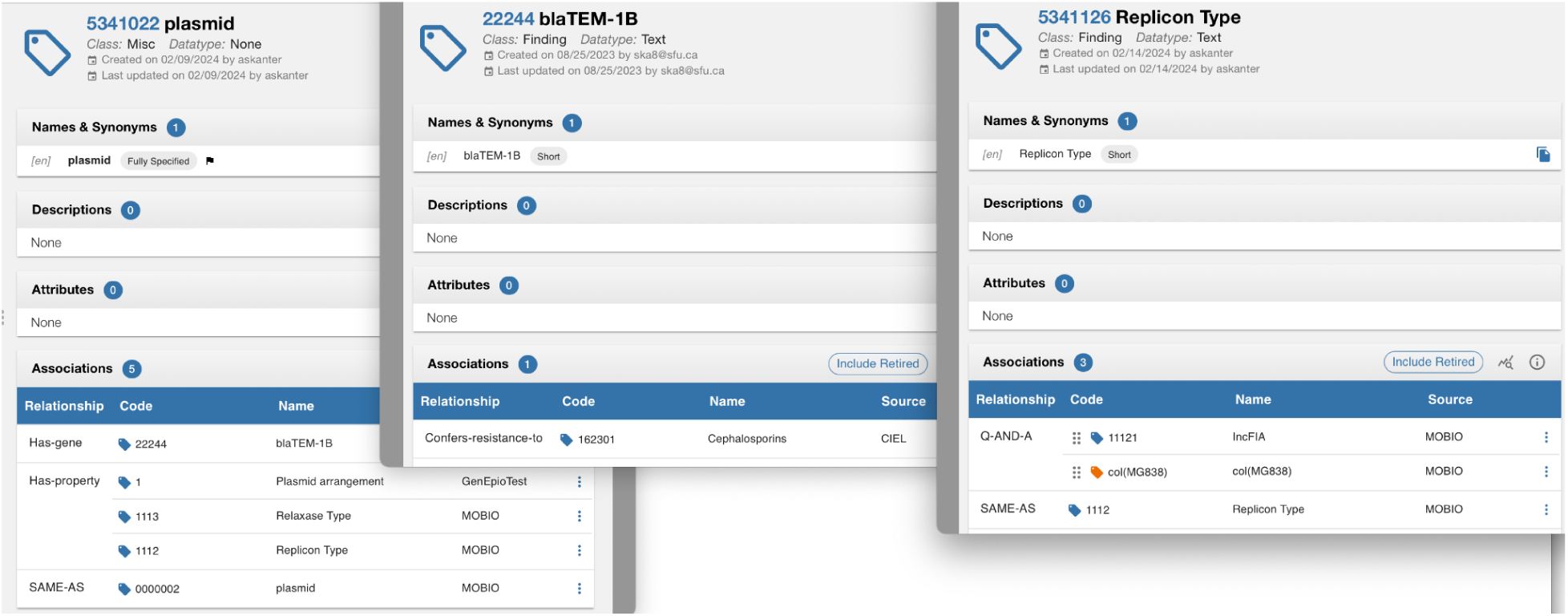
OCL screenshots showing mapping between the key concepts

**Table 2:**
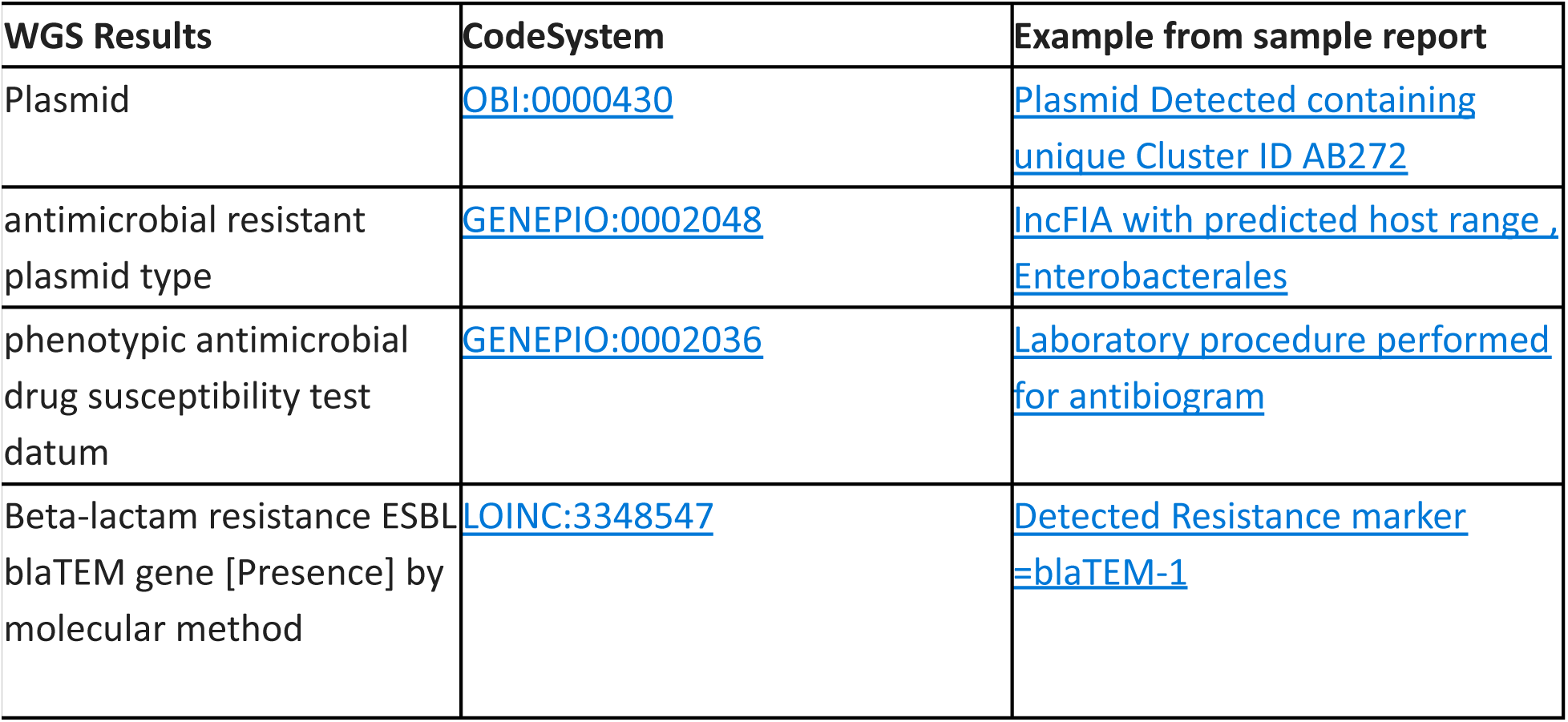
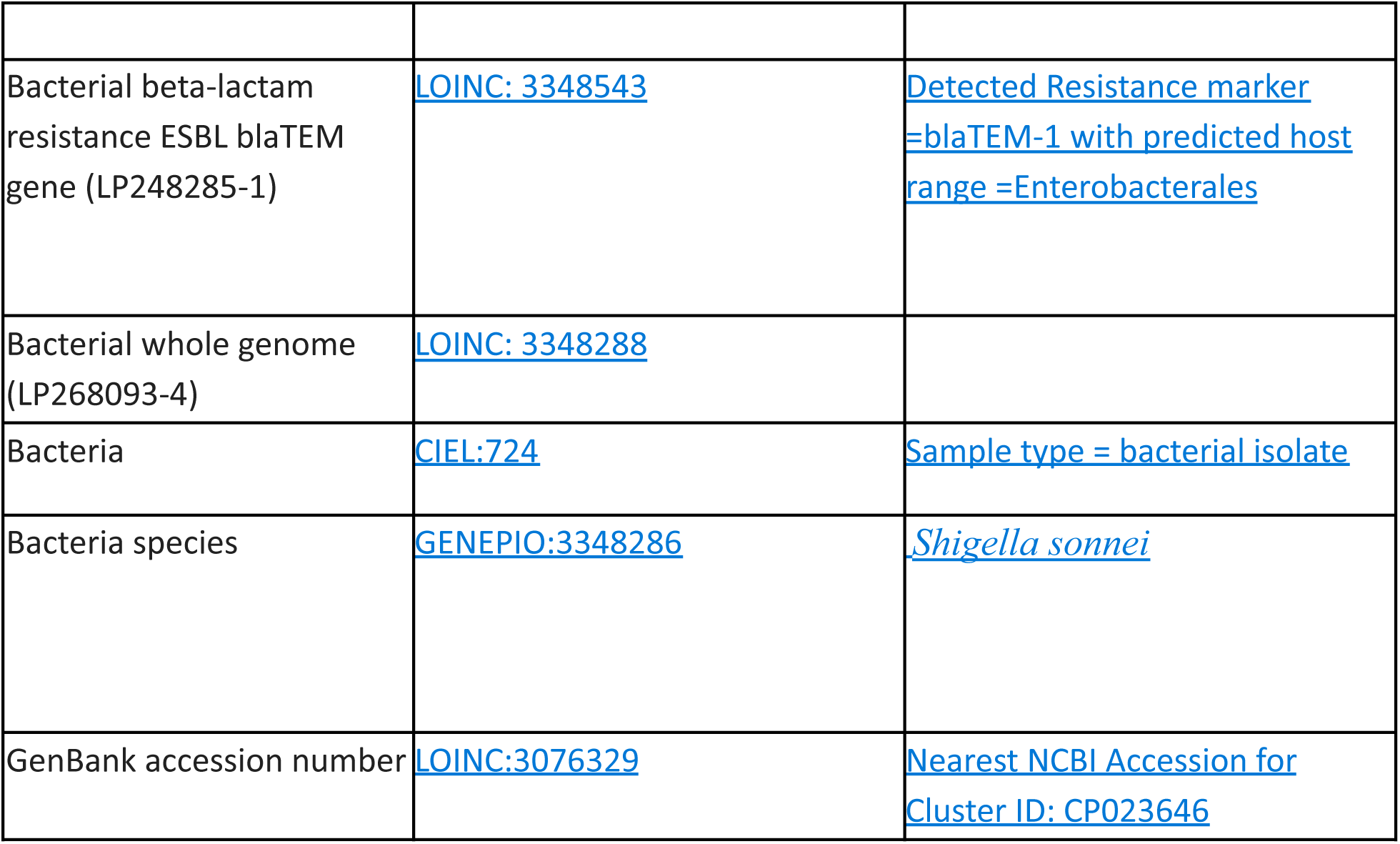
sample ValueSet Whole genome sequencing.

| WGS Results | CodeSystem | Example from sample report |
| --- | --- | --- |
| Plasmid | OBI:0000430 | Plasmid Detected containing unique Cluster ID AB272 |
| antimicrobial resistant plasmid type | GENEPIO:0002048 | IncFIA with predicted host range, Enterobacterales |
| phenotypic antimicrobial drug susceptibility test datum | GENEPIO:0002036 | Laboratory procedure performed for antibiogram |
| Beta-lactam resistance ESBL blaTEM gene [Presence] by molecular method | LOINC:3348547 | Detected Resistance marker =blaTEM-1 |
| Bacterial beta-lactam resistance ESBL blaTEM gene (LP248285-1) | LOINC: 3348543 | Detected Resistance marker =blaTEM-1 with predicted host range =Enterobacterales |
| Bacterial whole genome (LP268093-4) | LOINC: 3348288 |  |
| Bacteria | CIEL:724 | Sample type = bacterial isolate |
| Bacteria species | GENEPIO:3348286 | Shigella sonnei |
| GenBank accession number | LOINC:3076329 | Nearest NCBI Accession for Cluster ID: CP023646 |

Community-developed ontologies for the microbial genomics domain are made available in the OCL terminology server (https://openconceptlab.org/). These include the Antibiotic Resistance Ontology (ARO)(McArthur et al. 2013), which describes antibiotic resistance genes and mutations, their products, mechanisms, and associated phenotypes, as well as antibiotics and their molecular targets, and the MOBIO ontology (https://app.openconceptlab.org/#/orgs/cidgoh/sources/MOBIO/) which describes mobile genetic elements. GenEpio is an application ontology for genomic epidemiology, which covers four key areas including Food, Antimicrobial Resistance, Disease Surveillance and Mobile Elements.

Combining multiple ontologies as CodeSystems allows flexibility in development and encouages reuse. The aforementioned ontologies were selected for illustration purposes. For example, we used LOINC, the world’s most widely used terminology standard for health measurements, observations, and documents as well as the CIEL concept dictionary, which is a dictionary of over 53 thousand concepts relevant to health information systems in low- and middle-income countries (LMICs) whose concepts include diagnoses, procedures, medications, labs and observables (answers to questions) all mapped to international standard coding systems such as ICD-10-WHO, SNOMED CT, LOINC, RxNORM, and many others (https://app.openconceptlab.org/#/orgs/CIEL/). The CIEL concept dictionary was selected as it served as an interface terminology to work as a “bridge” between local terms and multiple reference and administrative codes.

Table 2 shows an example set of the ValueSet which contains key information associated with the whole genome sequencing result for a patient. The ValueSet is hosted within the OCL terminology server.

https://app.openconceptlab.org/#/orgs/cidgoh/sources/GenEpio/

Figure 6 demonstrates how Plasmid, blaTEM-1B, and Replicon Type concepts can map to other compatible ontologies (CodeSystems) shown as screenshots from OCL.

### FHIR data model for pathogen genomics data

Based on the review of the clinical scenario and review of CodeSystems and the information model, the following example demonstrates the data specification requirements for the FHIR observation resource. The whole genome sequence analysis can be represented using a profile of the observations resource. This new resource can consider:

1. Describe AMR genes and confer resistance
2. Identification of plasmid
3. Describe pathogen virulence factors such as toxic genes
4. Quality control metrics to indicate (signal quality index) sequencing quality
5. Should reference Patient
6. Should reference Practitioner
7. Can reference Encounter
8. Medication Adherence - MedicationDispense / MedicationStatement
9. ConferResistance may be treated like a MedicationDispense
10. MedicationAdministration might also be a resource we can use for medication adherence.
11. Make assumptions that the patient is prescribed with their medication from events like ConferResistance for MedicationStatement
12. References Patient
13. References Practitioner
14. Can use SNOMED CT medication code
15. Can reference Encounter, Procedure, Condition, Medication, Observation
16. Social determinant health Factors - Observations
17. Can generically describe almost anything related to the Patient’s environment (homeless, travel status,etc.)

### Scenario modelling for public health population data analysis aggregated at the point of care

This scenario involves a use case for bulk data access to be used for population-level analysis. The prior scenario involved a clinical setting for an individual patient. The working group discussed applications of expanded pathogen genomics knowledge from WGS for population-level analysis and how to best overcome the current limitations of data sharing. The scenarios involved bulk patient data access with the goal to allow standardized access by representing complex pathogen genomic data in a unified format, which includes both sequence data and the associated contextual data. No personal identifiable information was collected during the scenario modelling process. The use and integration of other apps and services such as 3rd party bioinformatics services were discussed as well as the suitability of FHIR tools given specific scenarios. To date, many 3rd party services are not FHIR compatible where only a few offers Application Programming Interface (API) integration. The privacy issue was also discussed regarding the use of social determinants of health data considering relational and structural aspects of infectious disease outbreaks affecting vulnerable communities. For example, Shigella infections disproportionally affect people experiencing homelessness.

An example of the scenario can be described as follows:

While investigating a patient with Shigella infection, WGS revealed the AMR genes are plasmid-born. AMR gene search found the same AMR genes in the Staphylococcus aureus infected patient within the same hospital under investigation for a hospital-acquired infection. Characterizing the plasmid across species of pathogens and the prevalence of the associated AMR genes are aggregated for regional and national reporting.

### Applying logic to structured pathogen genomic data

Computable clinical knowledge can be represented by numerous knowledge artifacts ranging from a simple relational query to a more complex bioinformatics process. For example, clinical decision support (“CDS”)(Mills 2019) is a well-established function within every hospital, which provides timely information both at the point of care and at population-level to inform relevant decisions for patient outcomes leveraging such computable clinical knowledge (i.e. logic used for clinical decision support and quality measurement). For example, "Patients with shigellosis should benefit from antimicrobial genotype profiling to identify risk factors predictive of treatment outcomes" is a statement of clinical knowledge.

In evaluating the computable clinical knowledge, we considered a scenario where there will soon be standardized and routine pathogen genomic investigations within the hospitals. Logic and FHIR operations are two relevant areas that provide foundations for clinical decision support based on genomic data. We used Clinical Quality Language (CQL), a high-level, domain-specific language focused on clinical quality as CQL can assist in building computable measures using FHIR resources and decision-support knowledge artifacts.

We experimented with CQL to create a library. We found the ease of creation, distribution, and execution of simplified computable knowledge based on experimental FHIR resources for pathogen genomic data information at the point of care. While building the library, we observed numerous benefits to using CQL as the healthcare-specific standard, which works out of the box with FHIR resources. Advantages include ease of development, mechanisms for verification, and the availability of tools to document and publish. CQL is data model independent, which facilitates automated execution and can work with ontologies based on new and emerging knowledge. The experimental IG containing the CQL library named “CQFPathogenGenomic” is available at (https://github.com/soyeangrey/CQFPathogenGenomic/). A screenshot of the IG artifacts (Figure 7) summary of local deployment is shown in Figure 7.

**Figure 7:**
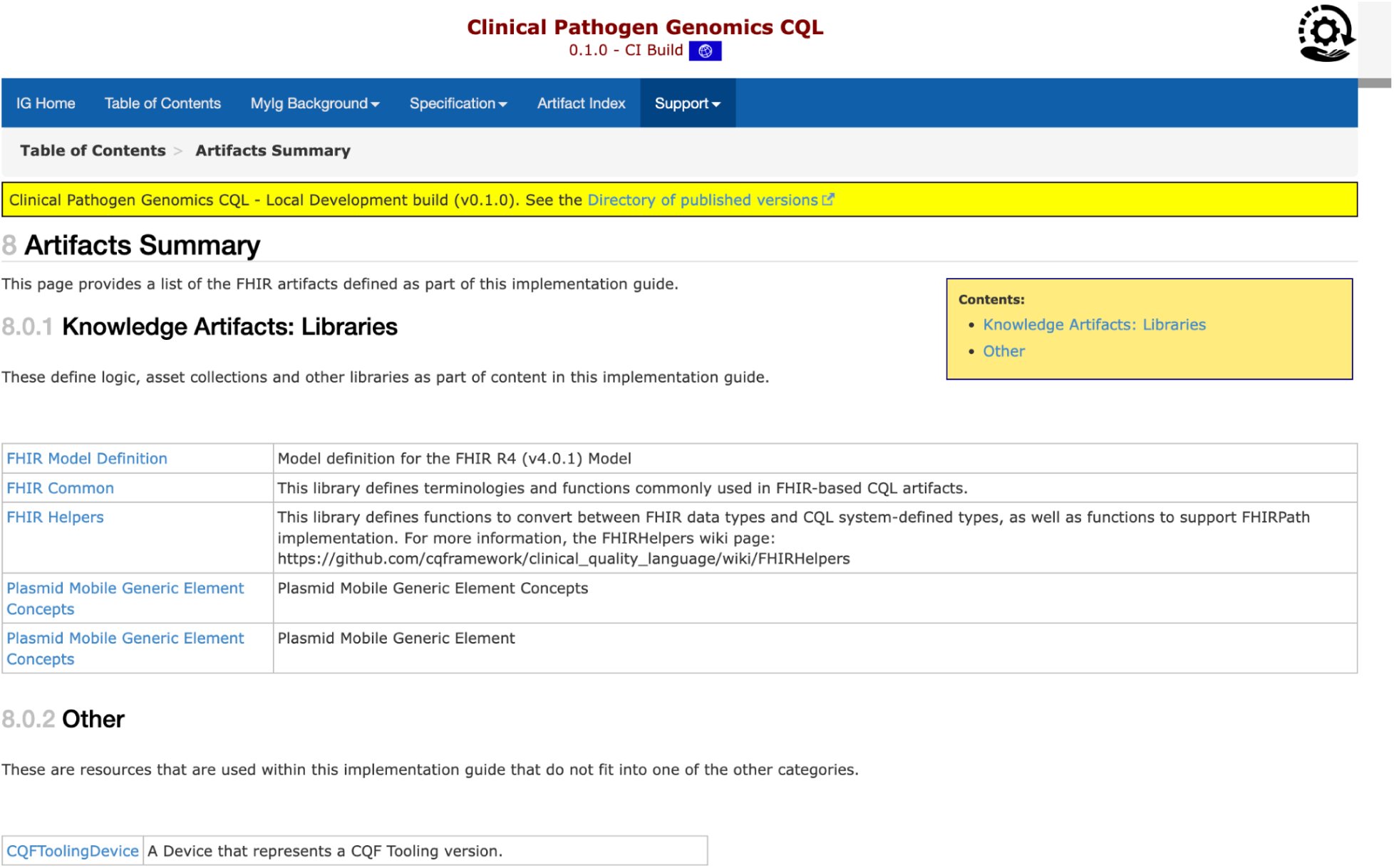
a screenshot of a demo IG

The ValueSets for the Clinical Pathogen Genomics IG are shown in Figure 8. Figure 8 contains experimental work only and does not have official URLs. The URLs are included here for illustration purposes only.

**Figure 8:**
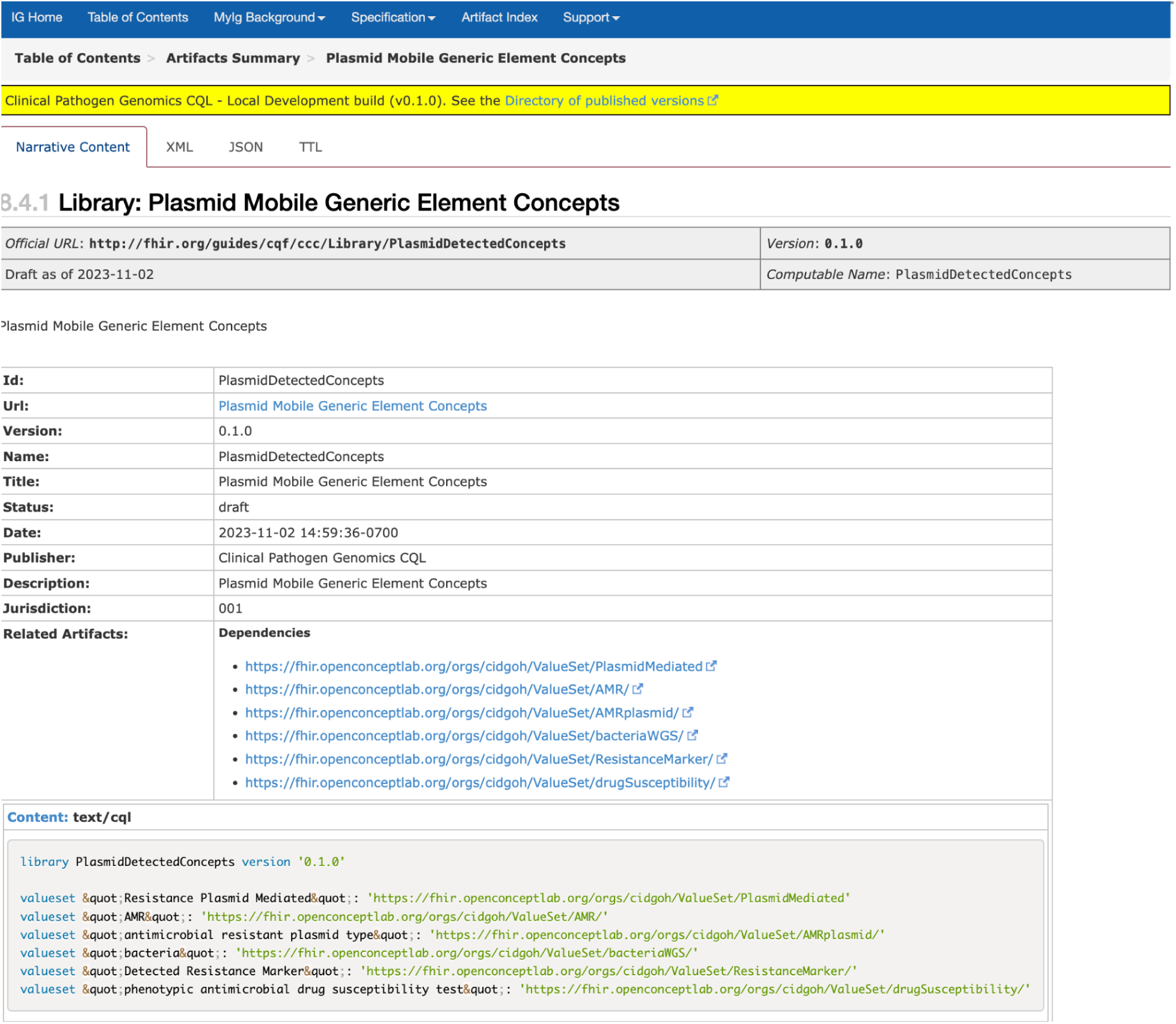
a screenshot of a demo IG showing key concepts

Test data was generated using a DiagonosticReport resource, which contains the code to indicate that the AMR gene found is plasmid-mediated (using the experimental CodeSystem and ValueSet published at OCL). Given the nature of the local outbreak, plasmid transmission was assumed. Figure 9 contains the recommendations as the result of executing the CQL library. This experiment was carried out using VSCode local runtime on a laptop (no public URLs). Figure 9 shows the result, which confirms the diagnostic report that references a patient in the test data containing the code indicating plasmid-mediated resistance. Based on that, the additional logic module in the CQL library is flagging this patient data for public health (“Is Flagged for Public Health” = true)

**Figure 9:**
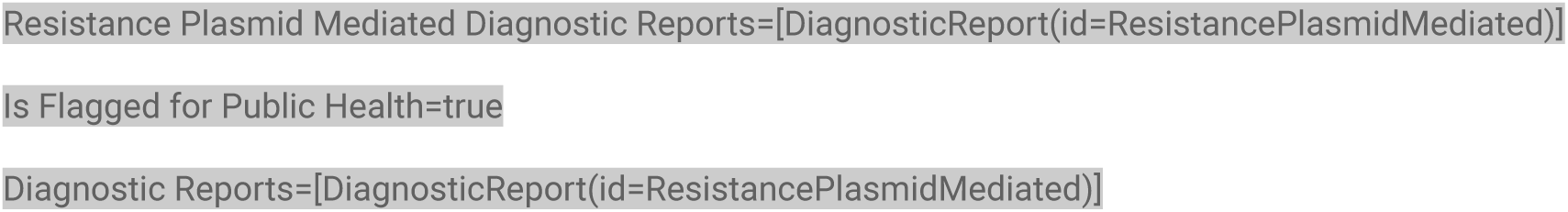
output from CQL library

### Managing ambiguity in clinical knowledge and genomic uncertainty

Genomic uncertainty (Ruscica et al. 2021) may arise when information from genomic testing is imperfect or unknown (i.e. no test result), leading to uncertainty in diagnosis or clinical decision support regarding optimal treatment. The uncertainty can come from many places such as patient encounters (i.e. incomplete data due to the patient being unconscious and therefore, unable to respond), testing, and/or throughout data management operation. This uncertainty due to ambiguity and complexity inherent to the information resulting from testing, quality of the sequence data, and bioinformatics processes (as well as the interpretations of the bioinformatics processes and results) must be managed.

In our experiment regarding the knowledge of whether resistance was plasmid mediated, if no exact match was found, several remediation strategies may be considered to deal with ambiguity in the genomic knowledge. This includes, but not limited to, using CQL to specify a conversion precedence for resolving the ambiguity. When matching the invocation type of an argument to the declared type of the corresponding argument of an operator, a compatible invocation type can be applied in addition to the exact match. In addition, an empty reason (i.e. ValueSet emptyReason) can be specified as a required element (if no match).

Well-known curated knowledge databases contain information regarding evolving pathogens and the annotation can change over time. The associated knowledge regarding intrinsic and plasmid-mediated organisms may change, which in turn, can return conflicting information and create ambiguity if the time of inference for AMR detection is not precisely defined in the knowledge artifacts. AMR detection is done through similarity to known sequences of antibiotic resistance genes(Alcock et al. 2020). Thus, a knowledge base can use AMR detection models to aid in scaling knowledge of treatment choices to previously unseen sequence variations. In addition to allowing CDS to scale, AMR sequence homology models(Yao and Yiu 2019; Alcock et al. 2020) will enable us to escape the combinatorial problem of nucleotide variation. Transitive annotation allows us to apply what we know about an annotated sequence to a novel sequence. Thus, it is key to include model matching in any knowledge base to reduce the exponential complexity of naming every species/variation combination. The use of similarity to organize knowledge can be seen in the CARD database where the degree of similarity between genome sequences can be expressed to reduce uncertainty. The CARD is a rigorously curated collection of characterized, peer-reviewed resistance determinants and associated antibiotics, organized by the Antibiotic Resistance Ontology (ARO) and AMR gene detection models (https://card.mcmaster.ca/).

### Conformance, Implementation guide (IG)

Implementation guides (IGs) play an important role in interoperability as they provide a computable statement about how FHIR resources and exchange can be used for a particular scenario (Lichtner et al. 2023). An implementation guide (IG) is a set of rules with associated documentation so that by using the IGs, implementers can demonstrate conformance against set standards.

We have created and published a pathogen genomic-specific IG to demonstrate how pathogen genomic-specific terminology can be used to evaluate a particular logic, which can inform the public health monitoring process concerning Plasmid-mediated AMR genes using a DiagnosticReport resource.

The experimental work published here for computable FHIR pathogen genomics tools is not considered FHIR conformant until further review. Future review will require broader community consultation as well as the balloting and voting process to involve the broader global HL7 FHIR communities including the HL7 Terminology Authority (HTA, https://confluence.hl7.org/display/TA).

## Discussions

As we worked to guide pathogen genomic data interoperability, we observed numerous advantages to using healthcare-specific standards such as FHIR and CQL. Advantages include convenient information models, mechanisms for verification, and the availability of tools, documentation and expertise to assist in development. We have created a collection of FHIR-compatible tools comprised of ontologies and IG, capable of supporting pathogen genomic use cases. This repository will assist in building the foundation to enable diverse, computable, and standardized pathogen genomic knowledge that will support automated analysis. In addition, the methods and tools we used to bring together various building blocks of interoperability are documented and can be adopted by other researchers and developers.

We also found the critical role of community-driven domain-specific ontologies which provide a wealth of knowledge addressing content coverage gaps in the common clinical terminologies (i.e. AMR genotypes and mobile genetic elements). While not fully formalized to integrate with the information model necessary for clinical operation, these community-vetted ontologies (i.e. OBO Foundry) provide foundational structures and semantics of emerging genomic knowledge, which can benefit from the proposed development paths and streamlined evaluation process (i.e. validation of ValueSets).

The proposed work lays the groundwork for CDS as an important stepping stone to scaling CDS based on Antimicrobial Sequence Detection (AMD) models. The use of AMD models overcomes the combinatorial explosion of sequence variation and complexity involved with the taxonomic resolution of bacterial communities. AMD models reduce the complexity of CDS rule-making. (e.g. replace the highly specific named resistance gene with the gene family).

We also experienced many challenges that others who are looking to implement new and complex knowledge as FHIR implementers may face. We encountered numerous occurrences of ambiguity and uncertainty arising from many places (from transitive annotation to date of inference). They include the choice of a curated knowledge database, empty data fields, sequencing quality and the time of inference.

Other challenges included balancing the need for simplicity (to allow ease of communication between clinicians and medical microbiologists) and complex semantics in the domain ontologies that originated from diverse processes and working towards clinically relevant knowledge artifacts without ambiguity. For example, there is a need for structure in the communication of drug susceptibility and resistance. MOBIO (Mobilome Ontology)(https://github.com/arpcard/mobio) describes both a higher-level concept for human consumption (such as gene family and bacterial species) and a lower-level, more precise representation of the variant sequence, along with important implications for a specific use case. The ambiguity (if not considered) on the terminology level sometimes resulted in a slowdown in implementation as they can affect downstream logical operations, causing translation issues in the narrative execution of the CQL library.

Finally, some genomic epidemiology concepts relied heavily on domain knowledge. This makes it difficult for non-clinicians or outsiders to replicate research or use existing concepts for new research. For example, ontologies such as ARO and MOBIO capture knowledge on the WGS test results, but there may be no information to connect the WGS result to other relevant information surrounding the WGS investigations. For example, reflex testing (or repeat/re-run) for a Diagnostic Report referencing the original test, which must be ordered first through ServiceRequest should contain an antibiogram that triggers a WGS investigation. Ontology curators should consider the surrounding context that may arise in clinical settings that need to be captured to be more precise at the terminology level.

Leveraging healthcare-specific standards such as FHIR and CQL comes with its learning curves, especially for non-health informatics professionals. However, we believe the potential benefits (speedy implementation and exchange of complex knowledge and reduced ambiguity) significantly outweigh the cost. We believe the proposed method is the fastest path to solving pathogen genomic data interoperability by allowing broad dissemination of the knowledge created by the whole community.

This work demonstrates a proof of concept to standardize computable genomic epidemiology workflow using point-of-care data. This includes human-readable and computable data formats and logic operations using genomic epidemiology domain ontologies and terminology binding. Given the benefits of real-time pathogen genomic data sharing for public health, we encourage public health researchers, ontology curators and implementors to use standardized computable definitions that can work with health care standards to allow integration of domain knowledge into clinical settings.

We acknowledge the limitations of this work. First, the tools developed here are limited in scope and not yet validated among the broader FHIR community. Therefore the ability to generalize for a broad set of pathogens is limited. Standardization of external ontology will require approval from the HL7 terminology authority. This approval process will require the demonstration of quality processes and measures and licensing and legal processes as well as community buy-ins. The information model here is developed based on scenario modelling. Additional validation using real clinical scenarios and patient data will be required for future developments. As the whole genome sequencing process is only beginning to emerge in clinical practices, more patient-derived WGS result data from multiple facilities will be needed to create generalized clinically valid pathogen genomic tools.

## Conclusion

To our knowledge, this is the first work of its kind to provide structured guidance on pathogen genomic data interoperability using HL7 FHIR resources for a clinical scenario involving whole genome sequencing. We believe this provides a clear path for broader stakeholders including implementors and knowledge curators on how to collaborate and facilitate automation in support of speedy exchange of complex knowledge for genomic epidemiology. We believe the tools and documentation provided can be a resource for clinical informatics, researchers, and public health organizations who want to collaborate, grow and exchange pathogen genomic knowledge for critical public health applications.

In the future, we plan to create a focused working group on pathogen genomics by working closely with the HL7 communities to accelerate the development and adoption of the HL7 FHIR as the standard to use computable pathogen genomic data products for public health research and patient care. We will initially focus on the evaluation and integration of standardized genomic epidemiology terminology (GenEpiO) to support real-time pathogen genomic data exchange using point-of-care information. Additional work will include profile development and incorporating Order and Observation resources as part of the IG. For example, building a transactional bundle to enable the loading of genomic reports with the labs into FHIR was out of the scope of this work, which may be considered in the future scope. Similarly, operations and queries to discover the sequence data (i.e. Molecular Sequence) together with structured findings (i.e. Observation) will also be considered. In reviewing existing IGs such as International Patient Summary (IPS)(“https://build.fhir.org/ig/HL7/fhir-ips/”), we see the potential to build a derivative of IPS by expanding the scope of the commonly reported pathogens (i.e. antibiogram profiles of antimicrobial susceptibility testing results). Future work will involve an in-depth review of the existing specifications in IPS, including data sharing mechanisms (i.e. patient consent) to explore standardized data access to both sequence data and key contextual information such as travel history and immunization records. The review will include how to leverage and re-profile existing Patient Summary (PS-CA) (https://simplifier.net/ps-ca-r1) to incorporate specialized profiles (i.e. Diagnostic Report containing WGS results) whose requirements may be added to the base level of constraints for reprofiling. The privacy issues surrounding the utilization of social determinants of health data (SDOH), while taking into account the relational and structural aspects of infectious disease outbreaks that impact vulnerable communities, will further require careful consideration prior to standardizing the discovery and access of SDOH data.

## Data Availability

All data produced in the present work are contained in the manuscript

## Acknowledgement

This work has been supported by grant to Dr. William Hsiao from the Canadian Institutes of Health Research (Project Grant Number: PJT-159456) and the Genome BC and Genome Canada (Grant Number: 286GET)

We gratefully acknowledge Dr. Daniel Holmes, Dr. Carolyn Shiau and Dr. Mel Krajden for their review and valuable suggestions.

